# Development of Chronic Pain and High-Impact Chronic Pain across the U.S. Rural-Urban Continuum, 2019-2020

**DOI:** 10.1101/2025.01.15.25320640

**Authors:** Feinuo Sun, Yulin Yang

## Abstract

**Purpose:** Rural health disadvantages are well-documented in previous literature, however research on rural-urban disparities in chronic pain outcomes is scarce. This paper fills this gap by examining pain prevalences and transitions across the rural-urban continuum (i.e., large central metro, large fringe metro, medium and small metro, and nonmetropolitan).

**Methods:** Based on the 2019-2020 National Health Interview Survey Longitudinal Cohort (NHIS-LC) data, we examined the disparities in pain prevalences and transitions among different pain states, including no pain, nonchronic pain, chronic pain and high-impact chronic pain (HICP), across the rural-urban continuum and by age, sex, race/ethnicity, and region. A test for linear trend was conducted to examine the significance of linear changes across the rural-urban continuum.

**Findings:** The findings reveal significant linear increases in the prevalence of chronic pain and HICP, as well as transitions from no pain to nonchronic pain and from nonchronic pain to more severe pain conditions, along the continuum from metropolitan to nonmetropolitan areas. Sub-group analyses indicate that rural-urban gaps are most pronounced among middle-aged (45-64) groups and non-Hispanic whites.

**Conclusions:** This study provides new evidence on rural-urban health disparities by focusing on pain, highlighting the urgent need to enhance healthcare services in remote and rural areas for effective pain prevention and management.

**Funding sources:** The authors received no external financial support for the research, authorship, or publication of this article.

**Disclosures:** The authors declare no potential conflicts of interest with respect to the research, authorship, or publication of this article.

## Introduction

Rural health disadvantages are well-documented across various health outcomes, largely due to the higher percentages of older and socioeconomically disadvantaged populations and limited healthcare services in rural areas.^1^ However, rural-urban disparities in pain, especially chronic pain (i.e., pain lasting more than 3 months), remain underexplored in existing research. This represents a significant gap, as chronic pain and high-impact chronic pain (i.e., HICP, chronic pain associated with limitations in daily living or work activities) have increasingly been recognized as pressing public health concerns in the U.S. given their rising prevalence over the past two decades^2,3^ and substantial health and economic consequences (e.g., increased risks of disability and mortality and healthcare costs).^3–5^ Only a limited number of studies indicate that rural residents experience worse pain outcomes compared to their urban and suburban counterparts,^6,7^ and little is known about how pain development and recovery differ across the rural-urban continuum. Utilizing the 2019-2020 National Health Interview Survey Longitudinal Cohort (NHIS-LC) data, this study provides the first analysis to evaluate the rural-urban disparities in transitions among different pain states—no pain, nonchronic pain, chronic pain, and HICP—and how these disparities vary across different populations.

## Methods

This cohort study uses the NHIS-LC data, including 10,415 respondents from the 2019 NHIS sample who agreed to participate in the 2020 follow-up. Initially there were 19,081 eligible participants, who were randomly selected from the 2019 sample, excluding those who were deceased, living in institutional settings, had proxy responses, or had missing contact information. Applying the longitudinal weights provided by NHIS, the sample represents the 2019 noninstitutionalized US civilian population.^8^ The University of Texas at Arlington Institutional Review Board judged this study is not considered as human subject research given it uses publicly available unidentifiable data. Analyses were conducted in November 2024.

Pain states were measured by two questions: “In the past three months, how often did you have pain, never, some days, most days, or every day?” and “Over the past three months, how often did pain limit your life or work activities, never, some days, most days, or every day?” Respondents reporting pain on “most days/every day” were categorized as having “chronic pain,” while pain on “some days” were coded as “nonchronic pain.” HICP referred to pain limiting life or work activities on “most days/every day.” We defined “pain incidence” as the transition from no pain in 2019 to nonchronic or chronic pain in 2020, “pain progression” as the transition from nonchronic pain to chronic pain, and “pain recovery” as the transition from chronic pain to nonchronic pain or no pain.^9^

Rural-urban continuum categories were based on the 2013 National Center for Health Statistics Urban-Rural Classification (i.e., the Office of Management and Budget’s definition) and included four ordered groups: “large central metro,” “large fringe metro,” “medium and small metro,” and “nonmetropolitan.”

We estimated pain prevalences and transitions across the rural-urban continuum and by age, sex, race/ethnicity, and region. A test for linear trend was conducted to examine the significance of linear changes across the rural-urban continuum.

## Results

The demographic characteristics of the NHIS-LC participants and those who did not participate are shown in the Appendix, with no significant differences found. According to Table 1, the weighted prevalences of any chronic pain and HICP in 2019 are 27.8% and 10.4%, respectively, in nonmetropolitan areas, significantly higher than those in large central metro areas (17.0% and 6.7%) and large fringe metro areas (17.7% and 5.6%). In contrast, the weighted prevalence of no pain is significantly lower in nonmetropolitan areas. The test for linear trend confirms that these differences are systematic across the rural-urban continuum. Table 2 further demonstrates that the percentages of transitions to nonchronic pain from no pain, to chronic pain or HICP from nonchronic pain, and to HICP from chronic pain all systematically increase from large metro areas to nonmetro areas over one year. Conversely, the percentages of individuals recovering from chronic pain to nonchronic pain or from nonchronic pain to no pain systematically decrease from large metro areas to nonmetro areas over one year.

**Table 1.** Weighted unadjusted prevalences with 95% confidence intervals and weighted frequencies of different pain status in 2019 across the rural-urban continuum for adult participants in the National Health Interview Survey Longitudinal Cohort

| **Pain status in 2019<sup>a</sup>** | **Large central metro** | **Large fringe metro** | **Medium and small metro** | **Non-metropolitan** | **p-value<sup>b</sup>** |
| --- | --- | --- | --- | --- | --- |
| **No pain** |  |  |  |  |  |
| % (95% CI) | 43.1 (40.7, 45.6) | 42.3 (39.5, 45.2) | 38.9 (36.7, 41.2) | 34.0 (30.9, 37.1) | <0.001 |
| Frequency | 1,417.9 | 1,039.8 | 1,234.0 | 506.5 |  |
| **Nonchronic pain** |  |  |  |  |  |
| % (95% CI) | 39.9 (37.6, 42.3) | 40.0 (37.2, 42.8) | 37.3 (35.2, 41.3) | 38.2 (35.2, 41.3) | 0.146 |
| Frequency | 1,311.3 | 981.2 | 1,184.3 | 570.4 |  |
| **Any chronic pain** |  |  |  |  |  |
| % (95% CI) | 17.0 (15.2, 18.9) | 17.7 (15.9, 19.7) | 23.8 (21.9, 25.7) | 27.8 (25.2, 30.6) | <0.001 |
| Frequency | 558.4 | 434.7 | 753.2 | 415.5 |  |
| **HICP** |  |  |  |  |  |
| % (95% CI) | 6.7 (5.6, 8.1) | 5.6 (4.5, 6.8) | 8.8 (7.6, 10.1) | 10.4 (8.7, 12.3) | <0.001 |
| Frequency | 220.3 | 136.5 | 277.4 | 154.8 |  |

CI: confidence interval; HICP: High-impact chronic pain
<sup>a</sup> Proportions of “no pain,” “nonchronic pain,” and “any chronic pain” are added up to 100.
<sup>b</sup> Results of the test for linear trend across the rural-urban continuum.

**Table 2.**
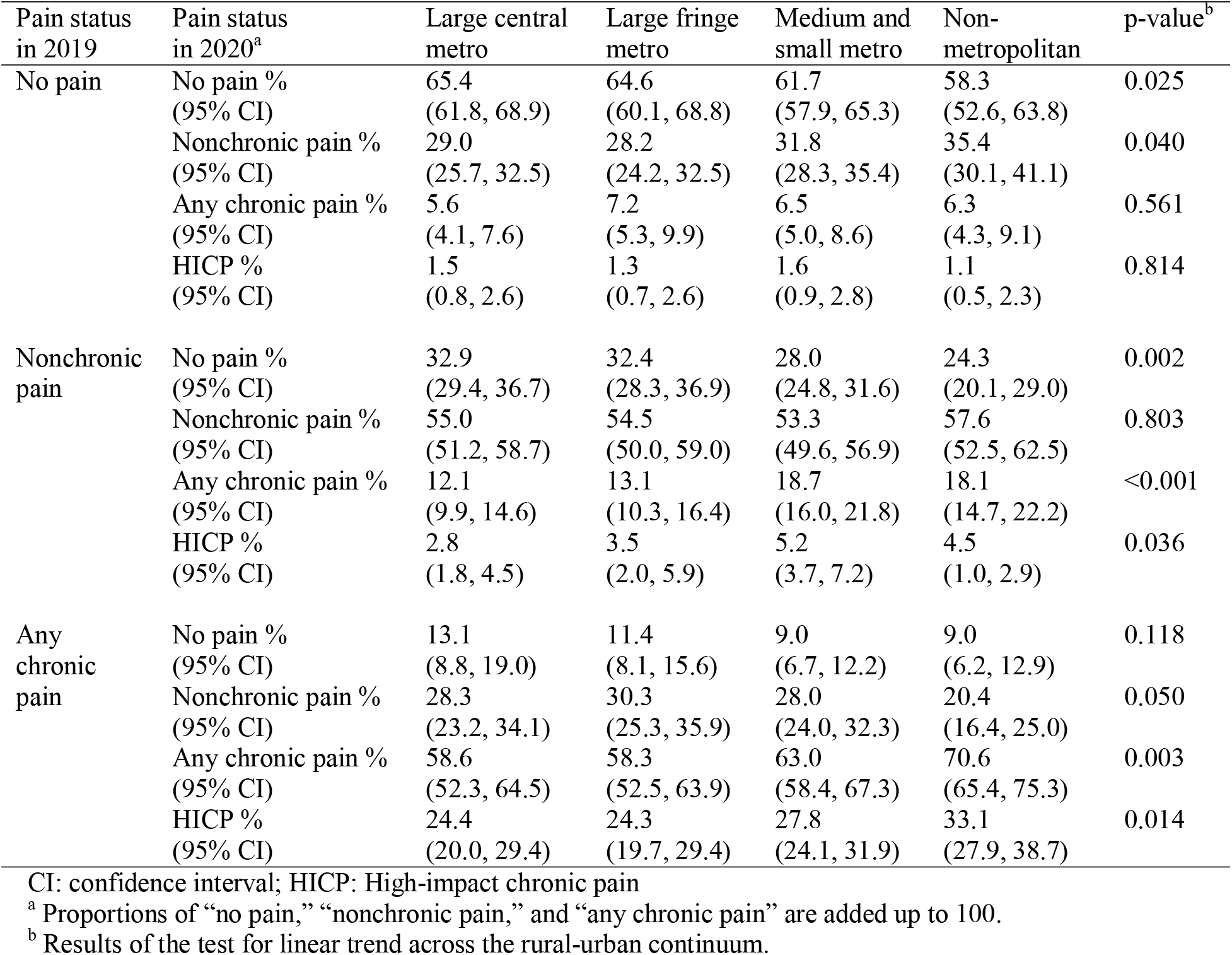
Weighted percentages of pain status with 95% confidence intervals in 2020 across the rural-urban continuum by pain status in 2019 for adult participants in the National Health Interview Survey Longitudinal Cohort

We further decomposed our analyses by demographic characteristics and census regions, shown in Table 3. Panels A, B, and C present the prevalences of pain incidence (baseline status: no pain), pain progression (baseline status: nonchronic pain), and pain recovery (baseline status: any chronic pain), respectively. For pain incidence, while rural-urban differences in estimates are not statistically significant, trend tests indicate a linear increase across the rural-urban continuum among middle-aged (45–64) individuals, males, and non-Hispanic White subgroups. The increase in pain progression across the rural-urban continuum is significant for most subgroups, including all age groups, both males and females, non-Hispanic Whites, and residents of the Northeast and West regions. Linear increases in recovery from chronic pain from more rural to more urban areas are observed only among middle-aged individuals (45–64 years old), females, and residents of the South.

**Table 3.** Weighted percentages of pain incidence, progression and recovery with 95% confidence intervals across the rural-urban continuum by demographic groups and regions for adult participants in the National Health Interview Survey Longitudinal Cohort

| A. 1-year cumulative incidence of any pain (95% CI), 2019-2020 |  |  |  |  |  |
| --- | --- | --- | --- | --- | --- |
|  | Large central metro | Large fringe metro | Medium and small metro | Nonmetropolitan | p-value<sup>a</sup> |
| Age |  |  |  |  |  |
| 18-44 | 32.8 (28.2, 37.8) | 32.0 (26.1, 38.6) | 34.7 (29.5, 40.3) | 36.8 (28.8, 45.7) | 0.387 |
| 45-64 | 35.1 (29.1, 41.7) | 39.4 (31.9, 47.5) | 43.6 (37.2, 50.2) | 48.0 (37.9, 58.3) | 0.017 |
| >65 | 41.9 (33.7, 50.6) | 40.8 (33.3, 48.9) | 41.8 (35.3, 48.5) | 46.7 (38.1, 55.5) | 0.559 |
| Gender |  |  |  |  |  |
| Male | 33.2 (28.5, 38.2) | 34.3 (28.6, 40.5) | 38.6 (33.4, 44.0) | 41.5 (33.4, 50.2) | 0.047 |
| Female | 36.1 (31.0, 41.5) | 36.5 (30.4, 43.1) | 38.0 (33.0, 43.3) | 41.9 (34.7, 49.4) | 0.252 |
| Race/ethnicity |  |  |  |  |  |
| NH White | 33.4 (28.9, 38.2) | 37.7 (32.5, 43.1) | 39.6 (35.4, 44.0) | 42.1 (35.9, 48.5) | 0.020 |
| NH Black | 39.6 (29.4, 50.9) | UR | 30.9 (19.8, 44.7) | UR | 0.505 |
| Hispanic | 34.0 (27.4, 41.4) | 31.6 (21.3, 44.0) | 38.2 (31.1, 50.6) | UR | 0.612 |
| Others | 33.8 (24.5, 44.5) | 24.2 (15.4, 35.8) | 30.8 (17.9, 47.6) | UR | 0.420 |
| Region |  |  |  |  |  |
| Northeast | 33.2 (25.2, 42.2) | 26.2 (19.5, 34.2) | 31.9 (23.7, 41.3) | UR | 0.963 |
| Midwest | 39.2 (30.8, 48.2) | 40.4 (32.0, 49.5) | 39.0 (31.2, 47.4) | 37.0 (29.1, 45.6) | 0.698 |
| South | 36.0 (29.7, 42.8) | 40.0 (32.4, 48.1) | 41.9 (36.3, 47.6) | 46.6 (37.5, 55.9) | 0.059 |
| West | 32.3 (26.7, 38.6) | 34.2 (24.4, 45.5) | 35.0 (27.6, 43.2) | UR | 0.329 |
| B. 1-year cumulative incidence of chronic pain from nonchronic pain (95% CI), 2019-2020 |  |  |  |  |  |
|  | Large central metro | Large fringe metro | Medium and small metro | Nonmetropolitan | p-value |
| Age |  |  |  |  |  |
| 18-44 | 8.8 (5.9, 12.9) | 6.7 (4.2, 10.7) | 14.8 (10.3, 20.7) | 12.3 (7.6, 19.2) | 0.050 |
| 45-64 | 15.6 (11.8, 20.4) | 16.2 (10.7, 23.9) | 21.8 (17.5, 26.8) | 22.2 (16.0, 30.1) | 0.032 |
| >65 | 13.6 (9.9, 18.5) | 19.7 (14.8, 25.7) | 21.5 (17.1, 26.6) | 20.8 (15.3, 27.7) | 0.028 |
| Gender |  |  |  |  |  |
| Male | 11.1 (8.2, 14.9) | 12.9 (9.0, 18.2) | 13.9 (10.8, 17.9) | 23.0 (17.2, 30.0) | 0.003 |
| Female | 12.9 (10.0, 16.5) | 13.2 (9.5, 17.9) | 23.0 (18.9, 27.7) | 14.4 (10.7, 18.9) | 0.014 |
| Race/ethnicity |  |  |  |  |  |
| NH White | 11.8 (9.1, 15.0) | 14.1 (11.1, 17.8) | 19.3 (16.5, 22.6) | 20.2 (16.3, 24.8) | <0.001 |
| NH Black | 15.1 (9.5, 23.1) | 8.6 (4.1, 17.0) | 24.6 (15.4, 36.8) | 11.2 (5.0, 23.2) | 0.431 |
| Hispanic | 13.4 (8.5, 20.5) | UR | 9.7 (4.6, 19.3) | UR | 0.370 |
| Others | 6.7 (3.2, 13.6) | 6.1 (1.1, 28.5) | UR | UR | 0.154 |
| Region |  |  |  |  |  |
| Northeast | 8.1 (4.8, 13.4) | 17.6 (10.3, 28.3) | 18.4 (12.8, 25.6) | 22.5 (11.3, 39.8) | 0.005 |
| Midwest | 13.8 (8.5, 21.5) | 11.2 (7.6, 16.3) | 15.8 (9.6, 24.9) | 20.1 (14.5, 27.2) | 0.100 |
| South | 14.1 (10.1, 19.4) | 12.1 (8.6, 16.9) | 20.3 (16.1, 25.3) | 14.0 (9.5, 20.2) | 0.237 |
| West | 11.4 (8.1, 15.7) | 10.9 (5.2, 21.3) | 18.4 (13.4, 24.7) | 22.2 (13.8, 33.6) | 0.009 |
| C. 1-year cumulative incidence of recovery from chronic pain (95% CI), 2019-2020 |  |  |  |  |  |
|  | Large central metro | Large fringe metro | Medium and small metro | Nonmetropolitan | p-value |
| Age |  |  |  |  |  |
| 18-44 | 55.0 (41.8, 67.5) | 56.6 (43.7, 68.6) | 51.8 (41.6, 61.9) | 33.8 (22.8, 46.9) | 0.058 |
| 45-64 | 33.9 (26.3, 42.3) | 34.0 (25.7, 43.3) | 25.5 (20.3, 31.4) | 24.7 (18.4, 32.2) | 0.025 |
| >65 | 36.5 (27.3, 46.9) | 40.8 (32.5, 49.7) | 36.9 (31.1, 43.0) | 31.7 (24.3, 40.2) | 0.347 |
| Gender |  |  |  |  |  |
| Male | 43.1 (34.5, 52.1) | 44.9 (36.2, 53.8) | 39.4 (32.6, 46.7) | 31.5 (24.6, 39.4) | 0.347 |
| Female | 40.1 (32.0, 48.9) | 38.9 (31.8, 46.6) | 35.1 (29.7, 40.9) | 27.5 (21.3, 34.8) | 0.029 |
| Race/ethnicity |  |  |  |  |  |
| NH White | 36.3 (29.6, 43.6) | 39.2 (33.1, 45.6) | 35.8 (31.0, 40.9) | 28.4 (23.4, 34.1) | 0.065 |
| NH Black | 43.6 (30.1, 58.1) | UR | UR | UR | 0.134 |
| Hispanic | UR | UR | 44.0 (29.9, 59.2) | UR | 0.494 |
| Others | UR | UR | UR | UR | 0.425 |
| Region |  |  |  |  |  |
| Northeast | 32.8 (20.9, 47.4) | 50.1 (38.9, 61.4) | 47.2 (33.9, 60.9) | 23.0 (11.9, 39.9) | 0.883 |
| Midwest | 49.1 (37.1, 61.1) | 33.9 (24.1, 45.2) | 33.8 (25.5, 43.4) | 34.3 (26.0, 43.8) | 0.075 |
| South | 41.3 (31.1, 52.3) | 37.9 (28.6, 48.2) | 35.8 (29.3, 42.9) | 26.4 (19.7, 34.5) | 0.016 |
| West | 42.1 (31.0, 54.0) | 48.4 (34.6, 62.6) | 35.2 (28.0, 43.1) | 33.4 (21.6, 47.8) | 0.194 |

CI: confidence interval; UR: Unreliable (does not meet National Center for Health Statistics standards for reliability)
<sup>a</sup> Results of the test for linear trend across the rural-urban continuum.

## Discussion

This study presents one of the first analyses of rural-urban disparities in pain and changes in pain states. Results show significant linear increases in prevalences of chronic pain and HICP in 2019, and percentages of people transitioning from milder pain conditions in 2019 to more severe ones in 2020 (e.g., from no pain to nonchronic pain or from nonchronic pain to chronic pain), from nonmetropolitan areas, the most rural, to large central metro areas, the most urban. Therefore, in addition to the higher pain prevalence in rural areas (e.g., the prevalence of any chronic pain in 2019 was over 10 percentage points higher in nonmetropolitan areas compared to large metro areas)—which may result from higher rates of chronic conditions and larger proportions of socioeconomic disadvantaged populations^1,10,11^—even when starting from the same baseline pain status, people in rural areas fare worse over one year compared to those in urban areas. This underscores a rural disadvantage in pain incidence and pain coping and management effectiveness, potentially due to healthcare shortages, limited access, and lower-quality services.

Rural-urban disparities in pain incidence and recovery rates are more pronounced for certain population groups than for others. Regarding pain incidence, these disparities are most notable among males, middle-aged adults, and non-Hispanic Whites—groups in rural areas particularly vulnerable to “deaths of despair.”^12^ Rural-urban gaps in pain recovery are significant among females, middle-aged adults, and residents in the South, indicating once pain develops, these groups in rural areas may face challenges in accessing effective approaches for managing and improving their conditions and thus need more attention.

The study has several limitations. First, our analysis only focuses on general self-reported pain without considering the underlying causes, specific locations, and durations of pain, which could be further explored in future research. Second, the percentages of transitions to severe pain conditions may be underestimated due to the healthy participant selection. Third, the small representation of racial/ethnic minority groups in rural areas and the low proportion of HICP cases makes some results unreliable and thus suppressed.

Nonetheless, this research contributes new evidence to the ongoing discussions on rural health disadvantages by focusing on pain. More efforts are needed to prevent pain incidence and improve the treatment and management of pain in rural areas, particularly among middle-aged individuals, to help close the rural-urban health gap.

## Data Availability

All data used in the present study are available publicly online at https://www.cdc.gov/nchs/nhis/documentation/2020-nhis.html
All Stata code is available on request to the authors.

## Notes

### Competing Interest Statement

The authors have declared no competing interest.

### Funding Statement

This study did not receive any funding

### Author Declarations

The study used publicly available data that were originally located at:https://www.cdc.gov/nchs/nhis/documentation/2020-nhis.html

### Summary of Updates

There is not significant change to the content, only the author list is updated.

