## Appendix Table for "Development of Chronic Pain and High-Impact Chronic Pain across the U.S. Rural-Urban Continuum, 2019-2020"

Appendix. Baseline characteristics of 2019 NHIS adult participants by enrollment status in the 2019-2020 longitudinal cohort

|  | 2019 enrolled  (longitudinal weights) | | | 2019 not enrolled  (sample adults weights) | | | Standardized difference scores | |
| --- | --- | --- | --- | --- | --- | --- | --- | --- |
| Variable | Raw Frequency | Weighted Frequency (1000's) | Weighted %  (95% CI) | Raw Frequency | Weighted Frequency (1000's) | Weighted %  (95% CI) | Un-weighted | Weighted |
| Total | 10,415 | 250,917 | 100 | 21,582 | 175,427 | 100 |  |  |
| Age |  |  |  |  |  |  |  |  |
| 18-44 | 3,441 | 115,265 | 45.9 (44.6, 47.3) | 8,583 | 84,198 | 48.0 (47.2, 48.8) | 0.141 | 0.042 |
| 45-64 | 3,569 | 82,655 | 32.9 (31.8, 34.1) | 7,028 | 56,366 | 32.1 (31.4, 32.9) | 0.036 | 0.017 |
| >65 | 3,405 | 52,996 | 21.1 (20.2, 22.0) | 5,971 | 34,863 | 19.9 (19.3, 20.5) | 0.109 | 0.030 |
| Gender |  |  |  |  |  |  |  |  |
| Male | 4,790 | 121,160 | 48.3 (47.0, 49.6) | 9,943 | 84,644 | 48.3 (47.4, 49.1) | 0.002 | 0.000 |
| Female | 5,624 | 129,741 | 51.7 (50.4, 53.0) | 11,637 | 90,770 | 51.7 (50.9, 52.6) | 0.002 | 0.000 |
| Unknown | 1 | 16 | UR | 2 | 13 | UR |  |  |
| Race/ethnicity |  |  |  |  |  |  |  |  |
| NH White | 1,153 | 41,506 | 16.5 (15.5, 17.7) | 2,999 | 30,722 | 17.5 (16.8, 18.2) | 0.085 | 0.027 |
| NH Black | 7,495 | 158,534 | 63.2 (61.8, 64.5) | 14,420 | 107,916 | 61.5 (60.7, 62.3) | 0.113 | 0.035 |
| Hispanic | 999 | 29,678 | 11.8 (10.9, 12.8) | 2,484 | 21,794 | 12.4 (11.9, 13.0) | 0.062 | 0.018 |
| Others | 768 | 21,198 | 8.4 (7.7, 9.2) | 1,679 | 14,995 | 8.5 (8.1, 9.0) | 0.015 | 0.004 |
| Region |  |  |  |  |  |  |  |  |
| Northeast | 1,789 | 44,572 | 17.8 (16.8, 18.8) | 3,621 | 31,140 | 17.8 (17.1, 18.4) | 0.011 | 0.000 |
| Midwest | 2,439 | 52,760 | 21.0 (20.0, 22.1) | 4,665 | 35,709 | 20.4 (19.7, 21.0) | 0.043 | 0.015 |
| South | 3,561 | 94,533 | 37.7 (36.4, 39.0) | 8,115 | 68,096 | 38.8 (38.0, 39.6) | 0.071 | 0.023 |
| West | 2,626 | 59,051 | 23.5 (22.4, 24.7) | 5,181 | 40,481 | 23.1 (22.4, 23.8) | 0.028 | 0.009 |
| Rural-urban continuum |  |  |  |  |  |  |  |  |
| Large central metro | 3,095 | 79,223 | 31.6 (30.3, 32.8) | 6,265 | 53,862 | 30.7 (30.0, 31.5) | 0.015 | 0.019 |
| Large fringe metro | 2,290 | 59,189 | 23.6 (22.5, 24.8) | 5,090 | 43,897 | 25.0 (24.3, 25.8) | 0.038 | 0.033 |
| Medium and small metro | 3,302 | 76,526 | 30.5 (29.3, 31.7) | 6,874 | 52,956 | 30.2 (29.5, 30.9) | 0.004 | 0.007 |
| Nonmetropolitan | 1,728 | 35,978 | 14.3 (13.5, 15.2) | 3,353 | 24,712 | 14.1 (13.6, 14.6) | 0.030 | 0.006 |

CI: confidence interval; UR: Unreliable (does not meet National Center for Health Statistics standards for reliability)
